# Predictive Capabilities of Polygenic Scores in an East-Asian Population-based Cohort: The Singapore Chinese Health Study

**DOI:** 10.1101/2025.02.13.25322249

**Authors:** Xuling Chang, Chih Chuan Shih, Jieqi Chen, Ai Shan Lee, Patrick Tan, Ling Wang, Jianjun Liu, Jingmei Li, Jian-Min Yuan, Chiea Chuen Khor, Woon-Puay Koh, Rajkumar Dorajoo

**Affiliations:** Department of Paediatrics, Yong Loo Lin School of Medicine, National University of Singapore, Singapore 119228, Singapore; Khoo Teck Puat – National University Children’s Medical Institute, National University Health System, Singapore 119074, Singapore; Department of Infectious Diseases, The University of Melbourne, The Peter Doherty Institute for Infection and Immunity, Melbourne, Australia, 3000; Genome Institute of Singapore (GIS), Agency for Science, Technology and Research (A*STAR), 60 Biopolis Street, Genome, Singapore 138672, Singapore; SingHealth Duke-NUS Institute of Precision Medicine, Singapore, Singapore; Cancer and Stem Cell Biology Program, Duke-NUS Medical School, Singapore, Singapore; Precision Health Research, Singapore, Singapore; Cancer Science Institute of Singapore, National University of Singapore, Singapore, Singapore; Department of Medicine, Yong Loo Lin School of Medicine, National University of Singapore, Singapore 117597, Singapore; Department of Surgery, Yong Loo Lin School of Medicine, National University of Singapore, Singapore 119228, Singapore; National Cancer Centre Singapore, Singapore, Singapore 168583; UPMC (University of Pittsburgh Medical Center) Hillman Cancer Center, Pittsburgh, PA 15232, USA; Department of Epidemiology, School of Public Health, University of Pittsburgh, Pittsburgh, PA 15261, USA; Singapore Eye Research Institute, Singapore National Eye Centre, Singapore 169856, Singapore; Healthy Longevity Translational Research Programme, Yong Loo Lin School of Medicine, National University of Singapore, Singapore 117545, Singapore; A*STAR Institute for Human Development and Potential (A*STAR IHDP), 30 Medical Drive, Brenner Centre for Molecular Medicine, Singapore 117609, Singapore

**Keywords:** polygenic scores, prediction ability, East-Asian

## Abstract

**Background:** Existing polygenic scores (PGS) are derived primarily from studies performed in European populations. It is still unclear how these perform in improving risk predictions in East-Asians.

**Methods:** We generated 2,173 PGSs from 519 traits and assessed their associations with 58 baseline phenotypes in the Singapore Chinese Health Study (SCHS), a prospective cohort of 23,622 middle-aged and older Chinese residing in Singapore. We used linear regression to evaluate PGS performances for quantitative traits by calculating the explained variance (r²). For dichotomized phenotypes, we employed logistic regression to estimate the area under the receiver operating characteristic curve (AUC) in predictive models.

**Results:** Overall, traits with higher heritability scores exhibited stronger associations with PGSs, while behavioural traits, for example sleep duration and hours spent watching TV, showed weaker associations. Height and type 2 diabetes (T2D) exhibited the largest SNP-based heritability estimates with the largest increments in explained variance and AUC, respectively, compared to phenotypic models. We explored the effect of T2D risk factors on the association between the T2D PGS (PGS003444) and incident T2D. The PGS association was significantly mediated and modified by hypertension (*P*_indirect_=1.56×10^−18^, *P*_interaction_=1.11×10^−6^) and body mass index (BMI, *P*_indirect_=1.25×10^−36^, *P*_interaction_=2.10×10^−3^). The prediction ability of PGS003444 for incident T2D was stronger was stronger among individuals who were non-overweight without hypertension (AUC=0.774) than in overweight individuals with hypertension (AUC=0.709).

**Conclusions:** In conclusion, our study demonstrated the divergent ability of PGSs in predictions of complex traits, and showed that for certain traits, such as T2D, PGSs may have the potential for improving risk prediction and personalized healthcare.

## Background

Progress in the field of genetics has significantly improved our understanding of the genetic basis of human diseases and complex traits (1). The introduction of genome-wide association study (GWAS) in the early 2000s has revolutionized genetic research by enabling the identification of thousands of genetic variants associated with various traits and diseases through the analysis of the entire genome in large populations (2). These discoveries have paved the way for the development and application of polygenic scores (PGSs), which aggregate the effects of numerous genetic variants and estimate an individual’s risk for specific traits or diseases based on their genomic profiles (3, 4). PGSs could serve in stratifying individuals depending on genetic predispositions and have been suggested as a cost-effective tool for population disease screening and for refining diagnostics and clinical interventions (5). Additionally, the emergence of valuable resources such as the PGS Catalog (6) has enabled researchers and clinicians to easily access and utilize PGSs, creating new opportunities for precision medicine and personalized treatments. The PGS Catalog is an open database of published PGSs, each of which has undergone standardized procedures and is consistently annotated with relevant metadata, including scoring files and details on the development and application of the PGS.

Despite significant progress in genetics research, disparities and global imbalances persist, as the majority of studies have been performed predominantly on European population samples. According to data from the GWAS Diversity Monitor (7), as of December 31, 2024, while 94.57% of GWAS participants are European, only 3.89 % are of Asian descent. Consequently, most PGSs have been developed based on GWAS data from European populations, which may hamper their transferability to other population groups (6). Genetic risk prediction accuracies of PGSs have been reported to be reduced in understudied population groups, including East-Asians, as compared to European population groups (8). A recent East-Asian study conducted in a hospital setting has highlighted that significant variabilities in predictive performance exist for PGSs across multiple traits (9). At the same time, however, for certain disease outcomes, such as breast cancer, PGS developed from primarily European genetic studies still show good predictive capabilities in Asian populations (10, 11). Hence, it is still not clear which PGSs have strong predictive capabilities in a population setting among East-Asians and how the genetic associations using existing PGSs can be used to guide improvements in risk predictions.

In this study, we evaluated the prediction performance of PGSs for various traits and diseases within a large East-Asian population-based cohort (N=23,622, 10,342 males and 13,280 females). Leveraging on the PGS catalog, which includes data on genetic risk variants and their corresponding weights, we systematically generated 2,173 PGSs for 519 traits. We then evaluated the association between these PGSs and 58 traits at baseline, in the Singapore Chinese Health Study (SCHS).

## Methods

### Study population and traits evaluated

The SCHS is a long-term prospective population-based cohort study focused on genetic and environmental determinants of cancer and other chronic diseases in Singapore (12). Between April 1993 and December 1998, a total of 63,257 participants (27,959 men and 35,298 women) aged 45 to 74 years were enrolled. Since April 1994, a random sample of 3% of participants were re-contacted for donation of blood/buccal cells and spot urine specimens. In January 2001, biospecimens collection was expanded to include all consenting participants, with approximately half of the cohort eventually contributing blood or buccal samples. SCHS was approved by the Institutional Review Board (IRB) at the National University of Singapore, and written informed consent was obtained from all study participants.

Details of the traits utilised in the study are presented in Supplemental Table 1. During the baseline interviews, participants were interviewed in-person with a structured questionnaire, gathering information on demographics, anthropometrics and lifestyle factors. Dietary information was collected using food frequency questionnaire (FFQ), which assessed usual intake over the previous 12 months of 165 items identified in a prior 24-h recall study to represent the major food and beverage items [27]. Descriptions of dietary scores created for the SCHS participants and utilized in this study have been previously reported (13, 14, 15, 16, 17, 18). The Alternative Healthy Eating Index (AHEI-2010) was created as an alternative measure of diet quality to identify future risk of diet-related chronic disease (19). The Mediterranean diet score was designed to examine the relationship between dietary habits of Mediterranean communities and chronic disease risk (20), with a modified version (aMed) utilized in the current study (21). The Dietary Approaches to Stop Hypertension (DASH) was initially designed for hypertension management (22). The meat–dim sum and vegetable–fruit–soy dietary scores were derived by principal components analysis (PCA) based on baseline dietary data (23). Dietary total antioxidant capacity was derived using two widely recognized scores of integrated dietary consumption of antioxidant nutrients, the Comprehensive Dietary Antioxidant Index (CDAI) and Vitamin C Equivalent Antioxidant Capacity (VCEAC) (16, 24).

During recruitment interviews, participants self-reported their height and weight, which were used to compute body mass index (BMI; kg/m^2^). BMI groups were defined based on cut-offs recommended for East-Asian populations – underweight (BMI < 18.5 kg/m^2^), normal weight (BMI between 18.5 kg/m^2^ to 23.0 kg/m^2^), overweight (BMI between 23.0 kg.m^2^ to 27.5 kg/m^2^) and obese (BMI > 27.5 kg/m^2^) (25). Hours spent watching television was obtained from questionnaire data in the SCHS (26). Participants’ sleep duration was evaluated using the question “On the average, during the last year, how many hours did you sleep in a day, including naps?”. Sleep was categorised as short sleep duration (defined as sleep duration of <6 hours), long sleep duration (defined as having >8 hours) and recommended sleep duration (maintaining 6 to 8 hours of sleep) (27). Cigarette smoking was categorized as never-smokers and ever smokers (28). Reproductive information was obtained at the baseline interview regarding the age at menarche, age at first livebirth, number of biological children, use of oral contraceptive and duration, age at menopause (if applicable), type of menopause (natural, surgery, radiotherapy or medication-induced) and use of hormonal replacement therapy [44]. Self-reported medical history on diabetes, hypertension, heart-attack and stroke were also collected by asking “Have you been told by a doctor to have any of these conditions?”.

Blood pressure, including systolic blood pressure (SBP) and diastolic blood pressure (DBP), were measured among the SCHS participants. SBP and DBP readings were taken at 3-minute intervals, and the average of these readings was used in the study (29). Mean arterial pressure (MAP = (2 × DBP + SBP) / 3) and pulse pressure (PP = SBP – DBP) were calculated. Telomere length measurements were made from blood leukocytes and was determined through a validated multiplex quantitative PCR assay, as previously described (30, 31). Metabolite biomarkers were quantified from plasma using the blood NMR biomarker platform developed by Nightingale Health Ltd and 28 clinically validated biomarkers were evaluated in the study.

Pulmonary tuberculosis (PTB) cases were ascertained via linkage with the National Tuberculosis Notification Registry (32). For longevity, we ascertained survival till 31 December 2020, through linkage with the nationwide registry of births and deaths, and compared individuals who were alive and above 85 or 90 years old with those who had died before 70 or 75 years old from natural causes. Ascertainment of incident type 2 diabetes was done by asking the participants for a history of physician-diagnosed diabetes at baseline and subsequent follow-up interviews, using the question: “Have you been told by a doctor that you have diabetes?” If the answer was “yes,” participants were also asked for the age at which they were first diagnosed. Participants were classified as incident type 2 diabetes cases if they reported developing diabetes between the baseline interview and subsequent follow-up interviews. The accuracy of self-reported diabetes had been estimated to be 98.8% in the SCHS (33).

### Genotyping and PGS generation

A total of 27,308 DNA samples were whole genome genotyped using the Illumina Global Screening Array (GSA), including 18,114 samples genotyped on v1.0 and 9,194 samples genotyped on v2.0. Detailed information on genotyping and quality control (QC) protocols has been previously published (31). After QC, SNP alleles were standardized to the forward strand and mapped to the GRCh38 reference genome. Additional autosomal SNPs were imputed using Minimac4 (version 1.0.0) (34) based on a local population-specific reference panel consisting of 9,770 whole-genome sequences of local Singaporean population samples from the SG10K initiative (SG10K Health) (35) on the Research Assets Provisioning and Tracking Online Repository (RAPTOR) (36).

For data security purpose, especially with sensitive human genetic data, all PGSs for SCHS were generated through RAPTOR, leveraging Amazon Web Services (AWS) HealthOmics. All data staged to HealthOmics are always encrypted both at rest and in-flight using custom keys. All PGS harmonized score files available prior to July 5th, 2023, from the PGS Catalog (https://www.pgscatalog.org/downloads/) were downloaded into a designated, restricted access S3 bucket as part of the application to host static input data. The pipeline required three input files: a file containing a list of S3 presigned URLs for input data files, the genome build (GRCh38) to be used for analysis, and specifications on total disk size. During the preprocessing phase of each run, the following intermediate files were staged into an isolated HealthOmics managed filesystem. These include the input file containing the list of S3 preassigned URLs for the input data files and all score files corresponding to the selected genome build specified in the input parameters.

During computation runs, all SCHS genotype files (PLINK format - BIM, BED and FAM files) were staged into the HealthOmics managed filesystem. These PLINK files were processed independently against each score file through the following steps: 1) Processed score files were created from the original PGS score file that included data on harmonized chromosome and position coordinates, effect alleles, other allele sand effect weights. 2) Processed score files were separated into biallelic variants and non-biallelic variants (monomorphic or multi-allelic). 3) For multi-allelic variants, matching information from the input SCHS BIM file was retrieved based on variant coordinates, with variants retained when there was a single match with the variant from the SCHS BIM’s effect/alternate allele and alignment with the variant’s effect allele. For biallelic variants, variants were retained when both alleles in the SCHS BIM file aligned with the variant’s alleles recorded in the score file. 4) Lastly, the --score function in PLINK was used to generate each PGS in the SCHS dataset using the processed score files. A total of 3,671 PGSs were generated successfully in the study. After excluding scores with low (<5)/high (>1M) number of SNPs, as well as those with a high SNP missing rate in SCHS (>25%), 2,173 scores remained available for downstream analysis. **Statistical analysis**

Genome-wide Complex Trait Analysis (GCTA) (37) was used to estimate SNP-based heritability for baseline quantitative traits and self-report diseases status, as disease prevalence is required for dichotomised phenotypes. Heritability was not estimated for blood metabolites due to the availability of measurements for only a small subset of the study participants (N = 1,504) and it is not recommended to run a GCTA-GREML analysis with a small sample (37).

All PGSs were normalized using rank-based inverse normalization (z-scores). Linear regression was applied to evaluate the performance of PGSs for quantitative traits, by calculating the explained variance (r²). For dichotomized phenotypes, logistic regression was employed and the area under the receiver operating characteristic curve (AUC) was calculated for evaluation. Previous study has shown that most models exhibited improved AUC values with the inclusion of additional covariates, such as PCs (9), which was also observed in our study (data not shown). For clarity, the significance of the improvement in predictive accuracy after the inclusion of the genetic information were calculated by comparing models that included genetic information and additional covariates (age, gender, PGS and the first three PCs) to those without genetic information (age and gender). The likelihood ratio test was performed to determine the significance for r^2^ using the R package lmtest. To evaluate the significance of the AUC change, Delong’s method was applied with R package, pROC (38). Additionally, the 95% confidence interval for AUC was calculated using the same package. The Cox proportional hazards model was utilized to investigate the association between PGS and incident T2D using the coxph package in R. Interaction analysis was performed by incorporating the interaction term into the model. Plots were generated with the R package ggplot2. All the above analyses were performed using R version 4.4.0 (39). Mediation analysis was conducted using the gSEM module in STATA (version15) (40), with the adjustment for age, sex and the first three PCs as covariates.

## Results

Supplemental Figure 1 presents the workflow for processing the data from the PGS Catalog. In brief, we downloaded PGS score files from the PGS Catalog (N = 3,683) (6) and successfully generated 3,671 scores using PLINK (41). After quality control, we assessed the association between 2,173 scores (Supplemental Table 2) and 19 quantitative traits, 28 blood metabolites and 11 dichotomized phenotypes (Supplemental Table 1). The associations were categorized as primary associations if the PGS was specifically designed for the outcome trait (for eg. PGS designed for height is associated with height), while they were classified as secondary associations if the PGS was originally developed for a different trait but showed an association with the outcome trait (for eg. PGS designed for blood pressure is associated with height).

### Primary associations for PGSs in the SCHS dataset

A total of 25 traits had corresponding trait-specific PGS available in the PGS catalog and all primary associations were statistically significantly (*P* ≤ 3.27×10^-4^, Supplemental Table 3, Figure 1). Metabolite data were available for a smaller subset of the study participants (N = 1,504). To minimize potential bias due to differences in sample size, results for blood metabolites were presented separately (Supplemental Figure 2).

**Figure 1:**
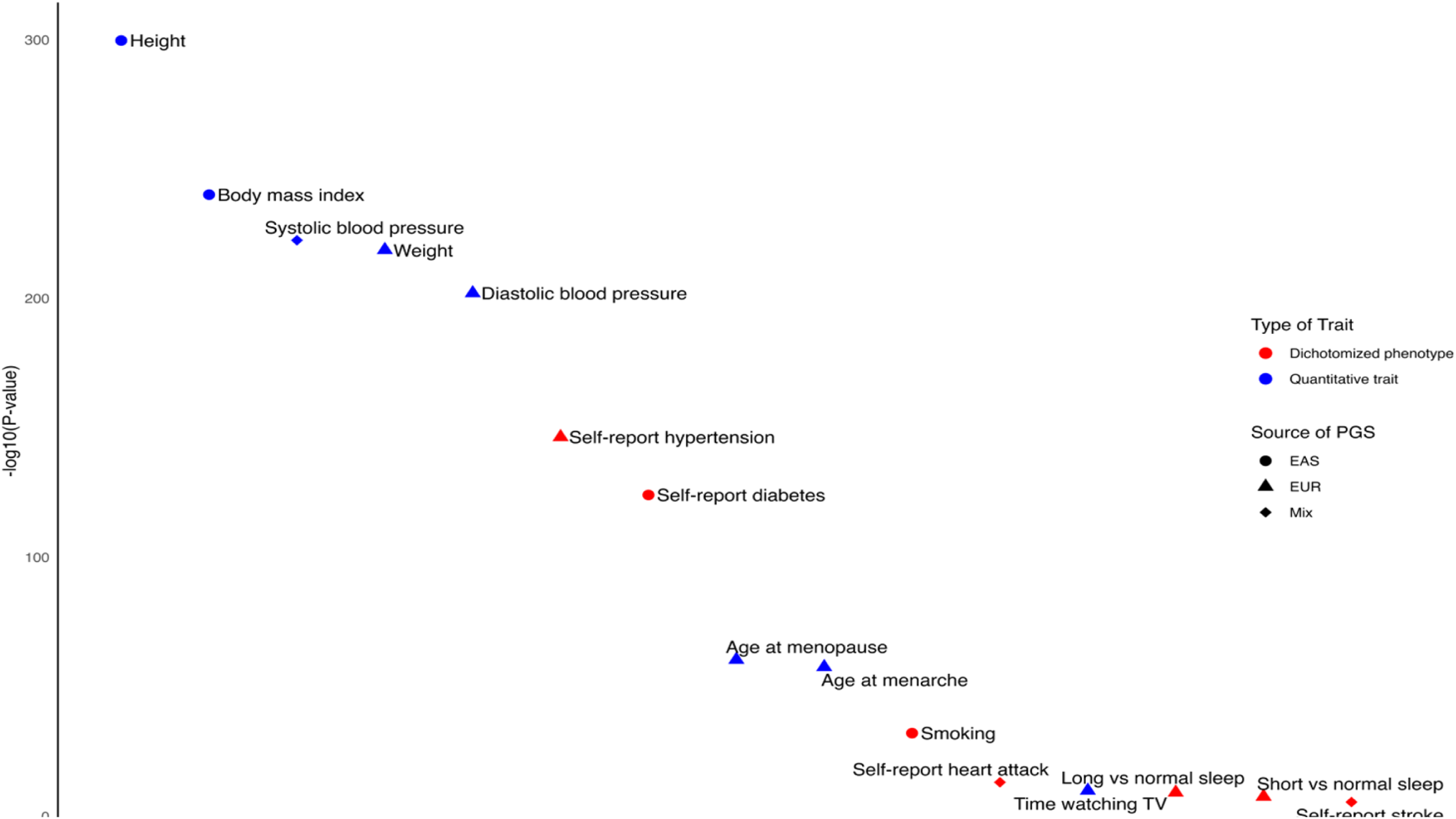
Primary PGS associations for baseline quantitative traits (blue) and dichotomized phenotypes (red) in SCHS. The shape of the dot indicated the source [source of variant associations (GWAS)]. If not available, information on the ancestry that the score developed/trained was employed. Circle: East Asian (EAS); Triangle: European; Diamond: Mixed population.

Heritability represents the theoretical upper limit of variance that can be explained by PGS (42). Among the quantitative traits, adult height exhibited the highest heritability estimate (h² = 0.472, Supplemental Table 3, Figure 2a). The most significant primary PGS associated with height with the largest effect size was PGS002803 (β = 0.295, *P* < 2×10^−300^), derived from GWAS genetic associations that were more relevant to East-Asians (43). Including PGS002803 in the model resulted in the greatest increase in explained variance (Δr^2^=0.089, Figure 2a). In contrast, behavioural traits such as time watching TV had the lowest heritability estimate (h² = 0.024, Supplemental Table 3, Figure 2a), the least significant primary association and the smallest effect size (β = 0.031, *P* = 1.19×10^−10^). Among the traits evaluated in the study, traits with higher SNP-based heritability generally showed greater explained variance in models after the incorporation of genetic information and more substantial increases in explained variance compared to models that only included phenotypic variables (Figure 2a).

**Figure 2:**
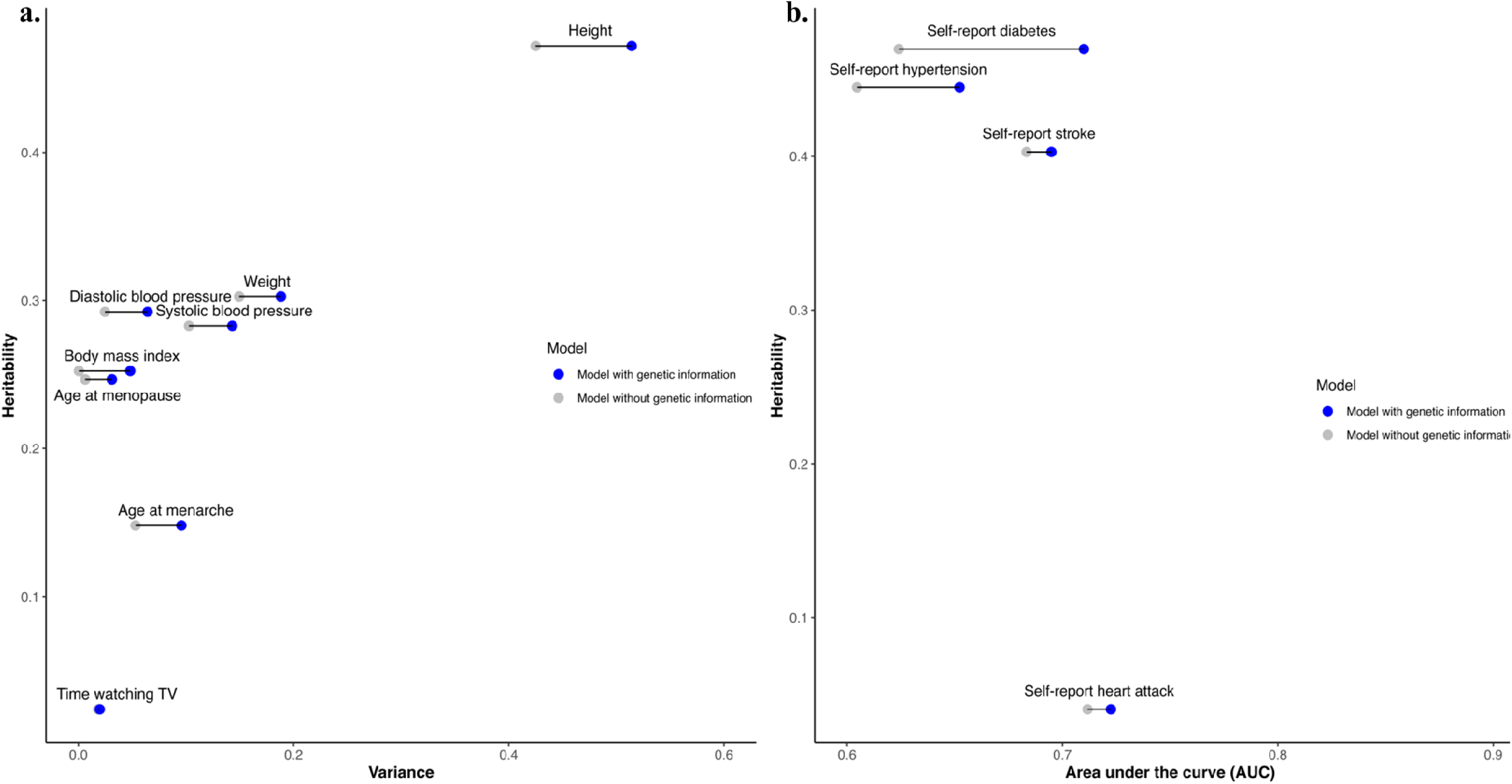
Relationship between trait heritability and predictive ability of primary PGS for **a.** quantitative traits; **b.** dichotomized phenotypes.

Among the blood metabolites, plasma high density lipoprotein (HDL) cholesterol exhibited the most significant association with the primary PGS (PGS002695, β = 0.286, *P* = 1.68×10^−36^, Supplemental Table 2) and showed the greatest increase in explained variance (Δr^2^=0.090, Supplemental Figure 2). While PGS002695 was derived from genetic associations from European population data, increments in r^2^ were noted to be consistent with previous evaluations among East-Asians (Δr^2^=0.102) and lower than that observed among European populations (Δr^2^=0.154) (44).

Among the binary traits evaluated in the study, type 2 diabetes (T2D) demonstrated the highest heritability estimate (h² = 0.470, Supplemental Table 3, Figure 2b). The most significant PGS associated with T2D was PGS003444, which was derived from a GWAS conducted in East Asian population samples (45, 46). While the association with T2D did not exhibit the highest predictive accuracy (AUC = 0.710), it showed the greatest increase in AUC when PGS003444 was included in the model (ΔAUC = 0.086, Supplemental Table 3). Consistent with the quantitative traits, binary outcomes with higher heritability generally exhibited a more pronounced increase in AUC upon incorporation of the PGS into the regression model, underscoring the varying significance of genetic contributions across traits (Figure 2b). Smoking status demonstrated the highest predictive accuracy in models both with (AUC = 0.837) and without (AUC = 0.831) genetic information (Supplemental Table 3). However, the PGS contributed only marginally to the predictive ability, suggesting that phenotypic variables such as age and gender accounted for a significantly larger proportion of the variance compared to genetic factors (ΔAUC = 0.006).

### Secondary associations for adult height and T2D

Given that adult height and T2D showed the highest SNP-based heritability estimates and the greatest improvements in predictive ability, we investigated their secondary associations in detail. Out of 12 PGSs for measurements commonly used to assess pulmonary function and respiratory health, 11 were significantly and consistently associated with adult height (*P* between 4.51×10^−164^ to 0.010, Supplemental Table 4). These include PGSs for vital capacity (EFO_0004312), forced expiratory volume (EFO_0004314), peak expiratory flow (EFO_0009718) and forced expiratory volume/forced vital capacity ratio (FEV/FVC Ratio; EFO_0004713), which are instrumental measurements in diagnosing respiratory conditions such as asthma and chronic obstructive pulmonary disease (COPD).

Another notable finding was the association between PGSs for liver-related enzymes and height in our dataset. PGSs for serum alanine aminotransferase (ALT) measurement (EFO_0004735) and aspartate aminotransferase (AST) to ALT ratio (AST/ALT ratio; EFO_0010934) were consistently associated with adult height (*P* between 5.49×10^−5^ and 0.002, Supplemental Table 4). The serum levels of these enzymes are commonly used to evaluate liver function and potential liver damage, including conditions like non-alcoholic fatty liver disease (NAFLD) (47), suggesting potential genetic links between adult height and liver function.

We also identified significant inverse associations between PGSs of traits such as sex hormone binding globulin (SHBG) (*P* between 9.58×10^−10^ and 5.79×10^−14^; Supplemental Table 5) and T2D risk. Significant associations were also observed between T2D and PGSs for anthropometric, endocrine and cardiometabolic traits, such as hypertension and BMI (Supplemental Table 5).

### Modification and mediation effects of hypertension and BMI categories on incident T2D

Previous studies have demonstrated that integrating genetic and environmental risk factors jointly into a model can enhance risk prediction for the risk of T2D (48). Guided by significant secondary pleiotropic PGS associations observed for T2D in our study, we evaluated how two key T2D risk factors, i.e. hypertension and BMI, could modify or mediate the relationship between the PGS and incident T2D. PGS003444 showed a significant association with incident T2D in our data [Hazard ratio (HR) (95%Cl) = 2.332 (2.243, 2.426), *P* < 2×10^−300^, Table 1]. Furthermore, HR for the association between PGS003444 and incident T2D decreased as BMI increased, with larger effects observed among underweight individuals [HR (95%Cl) = 2.770 (2.124, 3.612), *P* = 5.22×10^−14^, Table 1] as compared to the obese individuals [HR (95%Cl) = 1.947 (1.761, 2.154), *P* = 2.28×10^−38^, *P*_interaction_ = 1.11×10^−6^, Table 1, Supplemental Figure 3a]. Similarly, in stratified analysis by hypertension, the effect was stronger in the non-hypertensive group [HR (95%Cl) = 2.424 (2.310, 2.543), *P* = 3.75×10^− 287^, Table 1] than in those with hypertension [HR (95%Cl) = 2.112 (1.969, 2.266), *P* = 3.07×10^−97^, *P*_interaction_ = 2.10×10^−3^, Table 1, Supplemental Figure 3b]. These findings indicated that BMI and hypertension significantly modified the association between the PGS and incident T2D.

**Table 1:** Association between PGS003444 and incident type 2 diabetes stratified by body mass index groups or hypertension status.

| Group | case/non-cases | HR (95% CI) | *P* | *P*<sub>interaction</sub> |
| --- | --- | --- | --- | --- |
| All samples | 2625/18442 | 2.332 (2.243, 2.426) | < 2.00 x 10<sup>-300</sup> |  |
| Stratified by BMI groups |  |  |  |  |
| Underweight | 56/1299 | 2.770 (2.124, 3.612) | 5.22 x 10<sup>-14</sup> | 1.11 x 10<sup>-6</sup> |
| Normal weight | 752/8324 | 2.588 (2.411, 2.777) | 1.70 x 10<sup>-153</sup> |  |
| Overweight | 1366/7586 | 2.220 (2.100, 2.347) | 1.15 x 10<sup>-173</sup> |  |
| Obese | 451/1233 | 1.947 (1.761, 2.154) | 2.28 x 10<sup>-38</sup> |  |
| Stratified by hypertension status |  |  |  |  |
| Non-hypertensive | 1728/14909 | 2.424 (2.310, 2.543) | 3.75 x 10<sup>-287</sup> | 2.10x 10<sup>-3</sup> |
| Hypertensive | 895/3529 | 2.112 (1.969, 2.266) | 3.07 x 10<sup>-97</sup> |  |

In addition, we conducted a mediation analysis using Structural Equation Modeling (SEM), adjusting for age, sex and the first three principal components (PCs) as covariates. The analysis revealed that the effect of PGS on incident T2D was mediated by BMI (β_indirect-effect_ = 1.25×10^−36^) and hypertension (β_indirect-effect_ = 0.022, *P*_indirect-effect_ = 1.65×10^−18^, Supplemental Table 6).

Finally, we investigated the combined effect of overweight/obesity (BMI ≥ 23 kg/m) and hypertension on the risk predictive performance of PGS003444 on incident T2D. We first observed that the association was stronger in individuals who were neither overweight/obese nor hypertensive [HR (95%Cl) = 2.691 (2.484, 2.916), *P* = 6.83×10^−130^, Table 2, Figure 3a] than in individuals who were both overweight/obese and hypertensive [HR (95%Cl) = 2.024 (1.864, 2.199), *P* = 1.14×10^−62^, Table 2, Figure 3a]. The inclusion of PGS in the model significantly enhanced predictive performance, particularly for individuals who were non-overweight without hypertension, resulting in the highest AUC for models incorporating genetic information, with a notable increment in AUC (AUC = 0.774, ΔAUC = 0.227, Table 2, Figure 3b). In contrast, the T2D PGS had a lesser impact on individuals with both overweight/obesity and hypertension (AUC = 0.709, ΔAUC = 0.147, Table 2, Figure 3b).

**Table 2:**
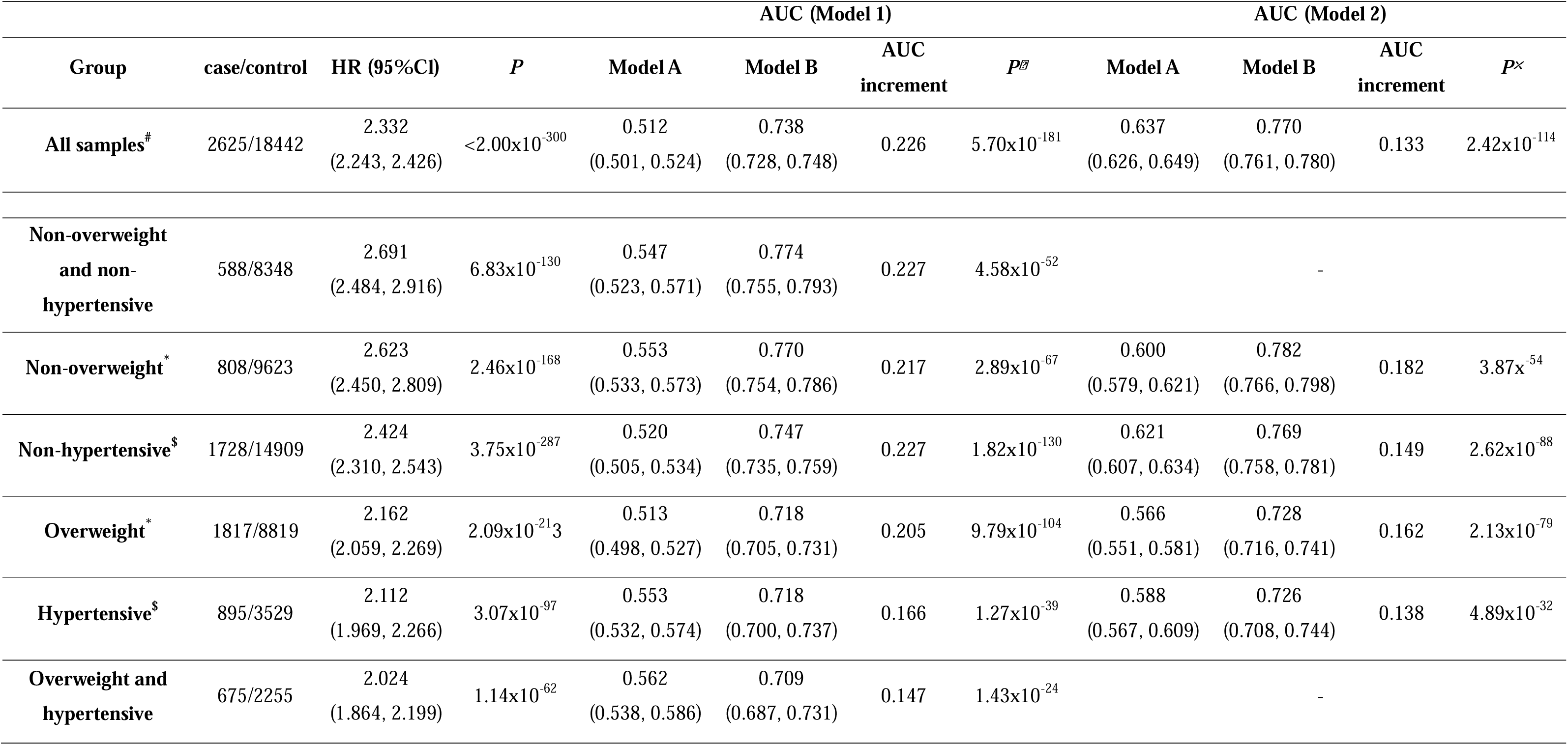
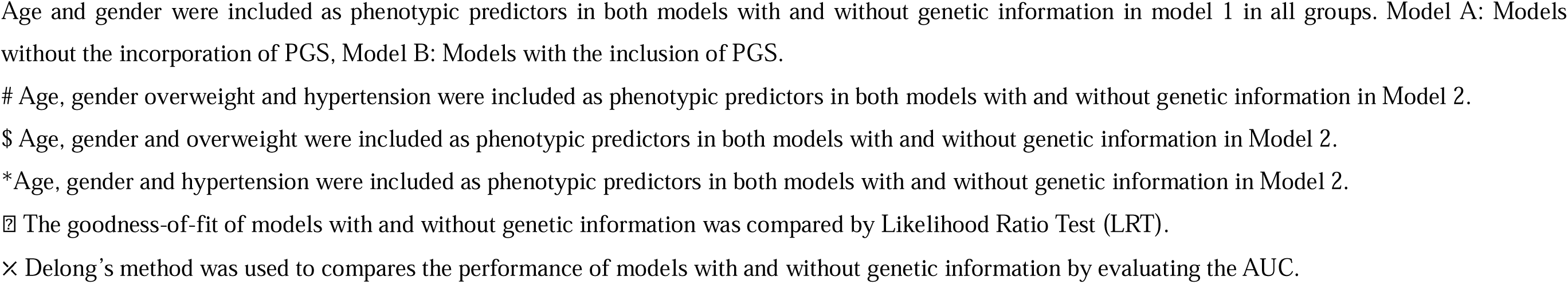
Association between PGS003444 and incident type 2 diabetes in SCHS stratified by overweight status, hypertension status and both risk factors.

| Group | case/control | HR (95%CI) | P | AUC (Model 1) |  |  |  | AUC (Model 2) |  |  |  |
| --- | --- | --- | --- | --- | --- | --- | --- | --- | --- | --- | --- |
|  |  |  |  | Model A | Model B | AUC increment | P<sup>§</sup> | Model A | Model B | AUC increment | P<sup>×</sup> |
| **All samples<sup>#</sup>** | 2625/18442 | 2.332<br>(2.243, 2.426) | <2.00x10<sup>-300</sup> | 0.512<br>(0.501, 0.524) | 0.738<br>(0.728, 0.748) | 0.226 | 5.70x10<sup>-181</sup> | 0.637<br>(0.626, 0.649) | 0.770<br>(0.761, 0.780) | 0.133 | 2.42x10<sup>-114</sup> |
| **Non-overweight**<br>**and non-**<br>**hypertensive** | 588/8348 | 2.691<br>(2.484, 2.916) | 6.83x10<sup>-130</sup> | 0.547<br>(0.523, 0.571) | 0.774<br>(0.755, 0.793) | 0.227 | 4.58x10<sup>-52</sup> |  | - |  |  |
| **Non-overweight<sup>\*</sup>** | 808/9623 | 2.623<br>(2.450, 2.809) | 2.46x10<sup>-168</sup> | 0.553<br>(0.533, 0.573) | 0.770<br>(0.754, 0.786) | 0.217 | 2.89x10<sup>-67</sup> | 0.600<br>(0.579, 0.621) | 0.782<br>(0.766, 0.798) | 0.182 | 3.87x<sup>-54</sup> |
| **Non-hypertensive<sup>\$</sup>** | 1728/14909 | 2.424<br>(2.310, 2.543) | 3.75x10<sup>-287</sup> | 0.520<br>(0.505, 0.534) | 0.747<br>(0.735, 0.759) | 0.227 | 1.82x10<sup>-130</sup> | 0.621<br>(0.607, 0.634) | 0.769<br>(0.758, 0.781) | 0.149 | 2.62x10<sup>-88</sup> |
| **Overweight<sup>\*</sup>** | 1817/8819 | 2.162<br>(2.059, 2.269) | 2.09x10<sup>-213</sup> | 0.513<br>(0.498, 0.527) | 0.718<br>(0.705, 0.731) | 0.205 | 9.79x10<sup>-104</sup> | 0.566<br>(0.551, 0.581) | 0.728<br>(0.716, 0.741) | 0.162 | 2.13x10<sup>-79</sup> |
| **Hypertensive<sup>\$</sup>** | 895/3529 | 2.112<br>(1.969, 2.266) | 3.07x10<sup>-97</sup> | 0.553<br>(0.532, 0.574) | 0.718<br>(0.700, 0.737) | 0.166 | 1.27x10<sup>-39</sup> | 0.588<br>(0.567, 0.609) | 0.726<br>(0.708, 0.744) | 0.138 | 4.89x10<sup>-32</sup> |
| **Overweight and**<br>**hypertensive** | 675/2255 | 2.024<br>(1.864, 2.199) | 1.14x10<sup>-62</sup> | 0.562<br>(0.538, 0.586) | 0.709<br>(0.687, 0.731) | 0.147 | 1.43x10<sup>-24</sup> |  | - |  |  |

Age and gender were included as phenotypic predictors in both models with and without genetic information in model 1 in all groups. Model A: Models without the incorporation of PGS, Model B: Models with the inclusion of PGS.

**Figure 3:**
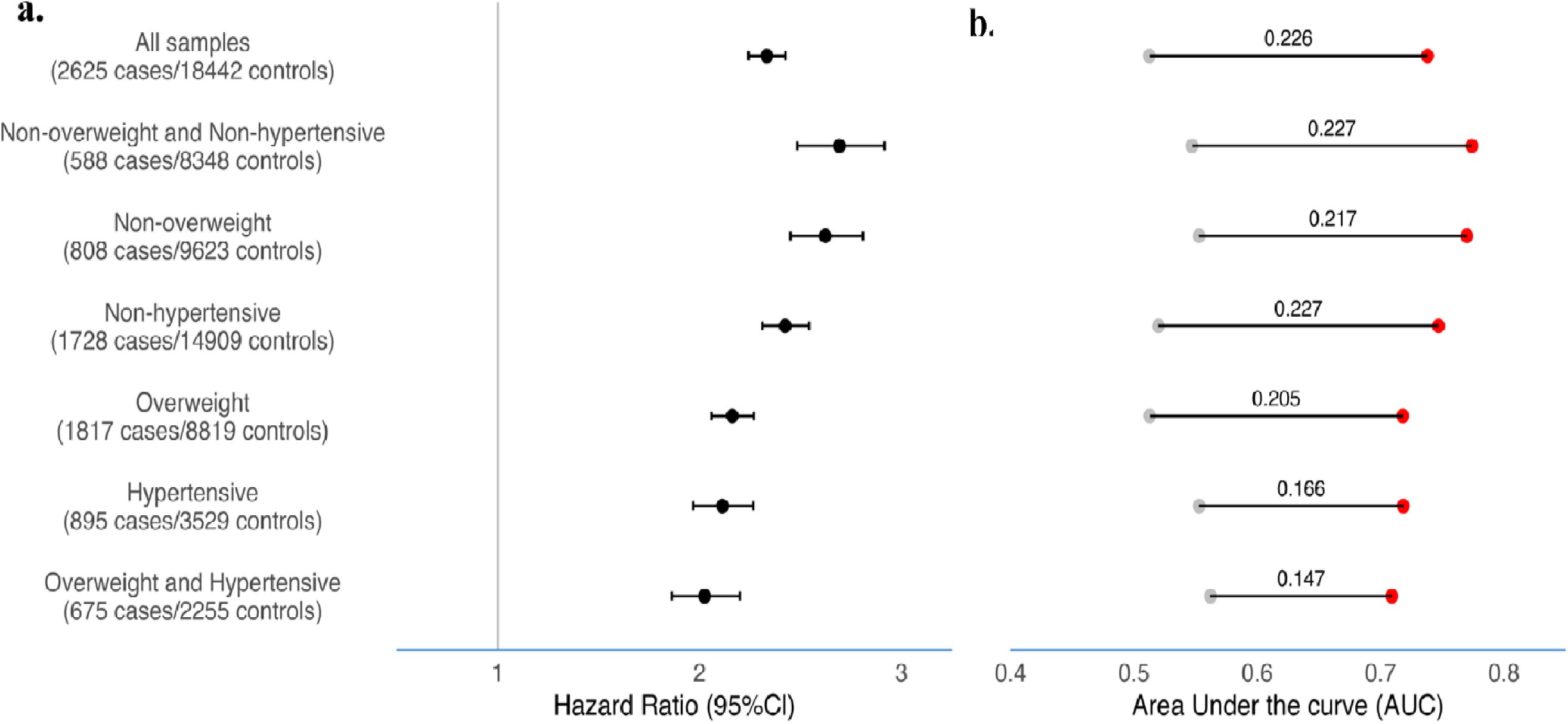
Association between PGS for type 2 diabetes mellitus (T2D; PGS003444) and incident T2D in SCHS stratified by obesity and hypertensive status. **a.** Hazard ratio and 95%Cl; **b.** the area under the receiver operating characteristic curve (AUC). The number above the line indicated the increment of AUC after inclusion on PGS003444 in the statistical models.

## Discussion

In this study, we conducted a systematic analysis of over 2,000 PGSs and evaluated their association with 58 baseline traits in an East-Asian population-based cohort. Our findings indicated that traits with higher heritability showed greater predictive improvements when genetic information was incorporated into the statistical models. Additionally, we observed that key risk factors, such as BMI and hypertension, could modify and mediate the effect of PGS on incident T2D. Furthermore, stratification using these risk factors, identified through PGS evaluations, has the potential to enhance risk prediction in the population.

A recent study in a hospital based setting among East-Asians revealed considerable variability in the predictive performance of PGSs across different disease traits, with AUC values ranging from 0.504 for oral aphthae to 0.874 for prostate cancer (9). Nearly 90% of traits exhibited AUCs within the 0.5–0.6 range, and the disease identification effectiveness of the PGS models largely fell within the AUC range of 0.6–0.7, underscoring challenges of applying PGSs derived from European populations to non-European populations. These findings were largely consistent with our study among the Chinese living in Singapore, with most of the PGSs effects on dichotomised phenotypes exhibiting AUC values between 0.5 to 0.7. We examined the origin of the PGSs and observed that among the 25 traits with primary association, five of the strongest PGS associations for these primary traits, including those for height and T2D, had PGSs derived from East-Asian genetic associations. Additionally, among traits where strongest associations were observed from PGSs derived from European-only genetic associations (such as PGS002695 for HDL levels), effect estimates were generally reduced in our East-Asian study. Taken together, these findings suggest that continued efforts are required to further identify population-specific genetic associations and to refine PGSs for improved predictive performances among Asian populations.

In our study, we extensively evaluated the secondary associations for height and self-reported T2D, as these traits exhibited the highest SNP-based heritability estimates and the greatest improvements in predictive performance. Genetic determinants for lung function were consistently associated with adult height in our study. Height has been identified as an independent risk factor for various respiratory diseases such as COPD and emphysema (49, 50). These secondary pleiotropic associations may facilitate further evaluations into the genetic connections between adult height and respiratory function. Significant associations were also observed between PGSs for blood enzymes commonly used to assess liver function and adult height. Elevated ALT/AST ratio is a recognised risk factor for liver dysfunction and development of NAFLD (51), while higher adult height has been associated with lower risk of NAFLD (52). For T2D, we identified significant associations between PGSs for SHBG and T2D. SHBG binds and transports androgens and estrogens in the blood, regulating their access to target tissues (53). Previous studies have reported that low serum concentrations of SHBG are associated with an increased risk of T2D (54, 55, 56). Consistently, Mendelian randomization studies have further suggested that serum levels of SHBG may have a causal effect on the risk of T2D (57, 58). Overall, the observed phenome-wide secondary associations between the PGSs and traits from our study reveal numerous genetic correlations between the traits evaluated. These genetic correlations warrant further evaluations in larger-scale independent genetic studies and systematic follow-up studies to improve understanding of disease mechanisms as well as to identify key risk factors that may serve in risk stratification and disease intervention strategies.

Guided by such secondary pleiotropic associations observed in our study, we demonstrated that genetically determined BMI and hypertension are significant risk factors for incident T2D. Our findings revealed that the strength of the association and the predictive accuracy of the T2D PGS on incident T2D were reduced in subjects with higher BMI. These results corroborated recent work (48). Data from BioBank Japan and UK Biobank found that the heritability for T2D was greater among individuals with lower BMI than those with higher BMI, and PGSs derived from low-BMI discovery outperformed PGSs from higher BMI levels (48). In the same way, we also found that hypertension significantly modified genetically determined predisposition for incident T2D. Furthermore, our analysis showed that the effect of the PGS on incident T2D was mediated through BMI and hypertension, and the predictive model performed best in non-overweight individuals without hypertension. These findings emphasize the importance of identifying factors that may modify the gene-disease association and thus affect disease risk predictions.

While the strengths of our study included detailed phenome-wide genetic evaluations in a homogenous East-Asian and well-curated population-based cohort, it is important to highlight potential limitations. First, the traits and diseases analysed were self-reported with questionnaires, which may introduce misclassifications due to erroneous replies from misremembering or misinterpretation of questions. Nonetheless, we observed significantly strong associations for all traits with primary PGSs, suggesting that the impact of such misclassification was likely minimal. Second, blood metabolite measurements were available for a smaller subset of participants, thus limiting the precision of SNP-based heritability estimates. Expanding these measurements to the full dataset would allow for a more comprehensive investigation into the relationship between PGSs and these metabolomics traits. Lastly, most of the PGSs were developed using European-derived summary statistics, which could diminish predictive accuracy when applied to non-European populations. To unlock the full and equitable potential of PGS, it would be essential to prioritize greater diversity in genetic studies.

## Conclusion

Our study provides a comprehensive evaluation of PGSs across a range of traits and diseases in a population-based Chinese cohort residing in Singapore, highlighting potential opportunities for precision medicine and personalized healthcare for certain disease outcomes. Additional secondary pleiotropic associations provide support for the potential of utilising existing PGSs to identify key factors for disease outcomes which, may serve to enhance disease prediction performance and identify subsets of at-risk individuals from the overall population.

## Data Availability

All polygenic risk scores used in this study are publicly available in the PGS Catalog (https://www.pgscatalog.org/). Significant associations between all PGSs and each of the 58 baseline traits evaluated in the study are detailed in Supplemental Table 7-62. Genetic information and generated PGSs for the SCHS are housed in the RAPTOR genomics repository (https://raptor.gisapps.org/). The data that support the findings of our study are available from the corresponding authors of the study upon reasonable request.

## List of abbreviations

ALT: Alanine aminotransferase
AHEI-2010: Alternative Healthy Eating Index
aMED: Alternate Mediterranean diet score
AWS: Amazon Web Services
AUC: Area under the receiver operating characteristic curve
AST: Aspartate aminotransferase
BMI: Body mass index
COPD: Chronic obstructive pulmonary disease
CDAI: Comprehensive Dietary Antioxidant Index
DBP: Diastolic blood pressure
DASH: Dietary Approaches to Stop Hypertension
FFQ: Food frequency questionnaire
GWAS: Genome-wide association study
GCTA: Genome-wide Complex Trait Analysis
GSA: Global Screening Array
HR: Hazard ratio
HDL: High density lipoprotein
IRB: Institutional Review Board
MAP: Mean arterial pressure
NAFLD: Non-alcoholic fatty liver disease
PGS: Polygenic scores
PCA: Principal components analysis
PTB: Pulmonary tuberculosis
PP: Pulse pressure
QC: Quality control
SHBG: Sex hormone binding globulin
SCHS: Singapore Chinese Health Study
SEM: Structural Equation Modeling
SBP: Systolic blood pressure
T2D: Type 2 diabetes
VCEAC: Vitamin C Equivalent Antioxidant Capacity

## Declarations

### Ethics approval and consent to participate

SCHS was approved by the Institutional Review Board (IRB) at the National University of Singapore, and written informed consent was obtained from all study participants.

### Availability of data and materials

All polygenic risk scores used in this study are publicly available in the PGS Catalog (https://www.pgscatalog.org/). Significant associations between all PGSs and each of the 58 baseline traits evaluated in the study are detailed in Supplemental Table 7-62. Genetic information and generated PGSs for the SCHS are housed in the RAPTOR genomics repository [51] (https://raptor.gisapps.org/). The data that support the findings of our study are available from the corresponding authors of the study upon reasonable request.

### Authors’ contributions

R.D., X.C., and W-P.K., contributed to the study design. W-P.K., and J-M.Y., contributed to the sample recruitment and data processing. C-C.K., and W-P.K., generated genotyping data. R.D., X.C., L.W., and C-C.K., performed genotype quality controls, imputation and genotyping analyses. R.D., X.C., L.W., and J.L., performed statistical analyses. R.D., C.C.H, J.C, A.S.L, P.T, J.L. and X.C., developed methodologies for data linkages and PGS generation. R.D., and X.C., verified underlying data. R.D. and X.C., drafted the manuscript. All authors read, critically reviewed and approved the final manuscript. R.D and X.C. are the guarantors of this work.

## Acknowledgements

We thank Siew-Hong Low of the National University of Singapore for supervising the fieldwork in the Singapore Chinese Health Study.

