## Supplemental Figures for "Predictive Capabilities of Polygenic Scores in an East-Asian Population-based Cohort: The Singapore Chinese Health Study"

### Slide 1
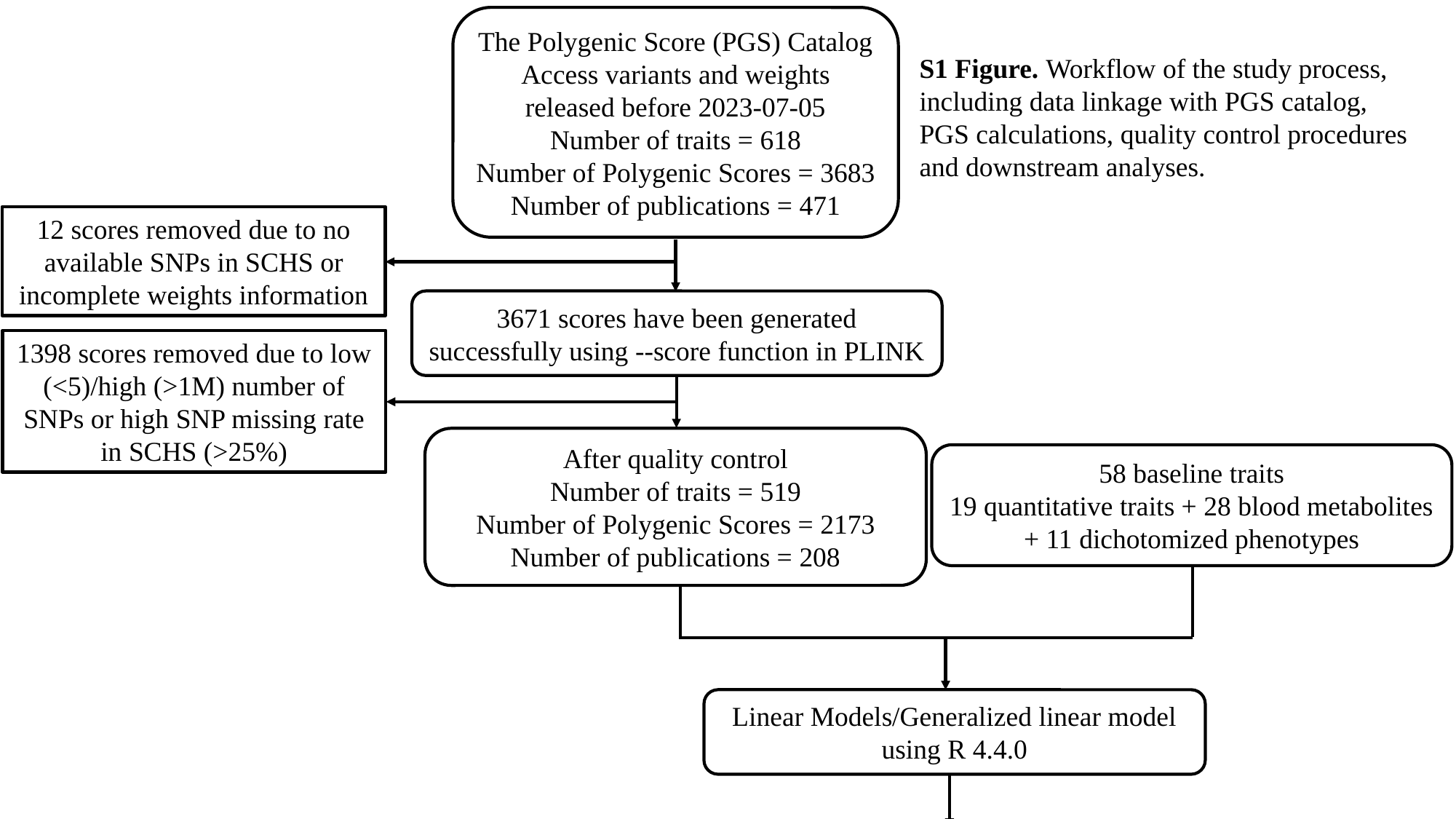

The Polygenic Score (PGS) Catalog
Access variants and weights released before 2023-07-05
Number of traits = 618
Number of Polygenic Scores = 3683
Number of publications = 471
S1 Figure. Workflow of the study process, including data linkage with PGS catalog, PGS calculations, quality control procedures and downstream analyses.
12 scores removed due to no available SNPs in SCHS or incomplete weights information
3671 scores have been generated successfully using --score function in PLINK
1398 scores removed due to low (<5)/high (>1M) number of SNPs or high SNP missing rate in SCHS (>25%)
After quality control
Number of traits = 519
Number of Polygenic Scores = 2173
Number of publications = 208
58 baseline traits
19 quantitative traits + 28 blood metabolites
+ 11 dichotomized phenotypes
Linear Models/Generalized linear model using R 4.4.0
Secondary analysis: Mediation/Modification

### Slide 2
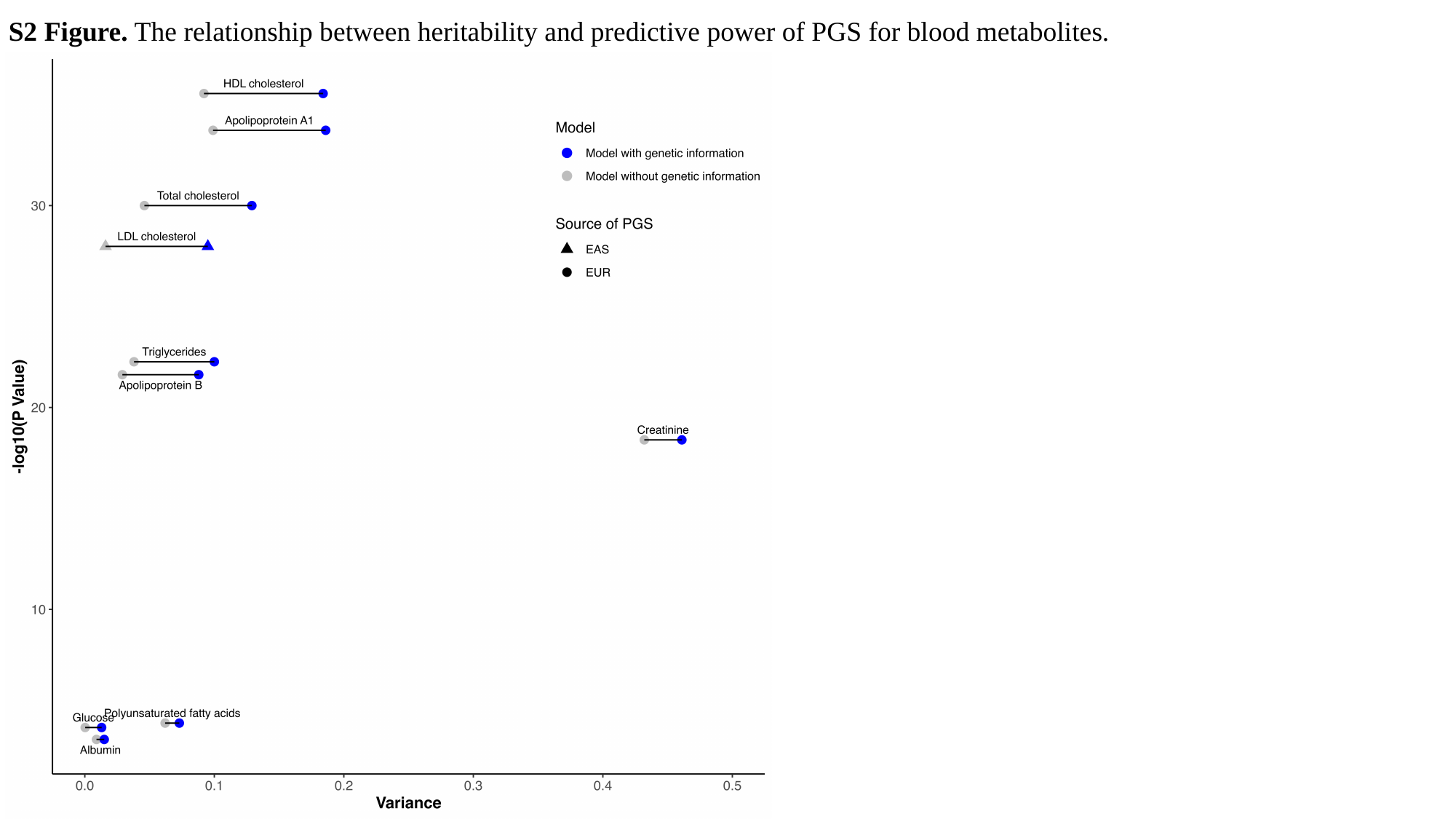

S2 Figure. The relationship between heritability and predictive power of PGS for blood metabolites.

### Slide 3
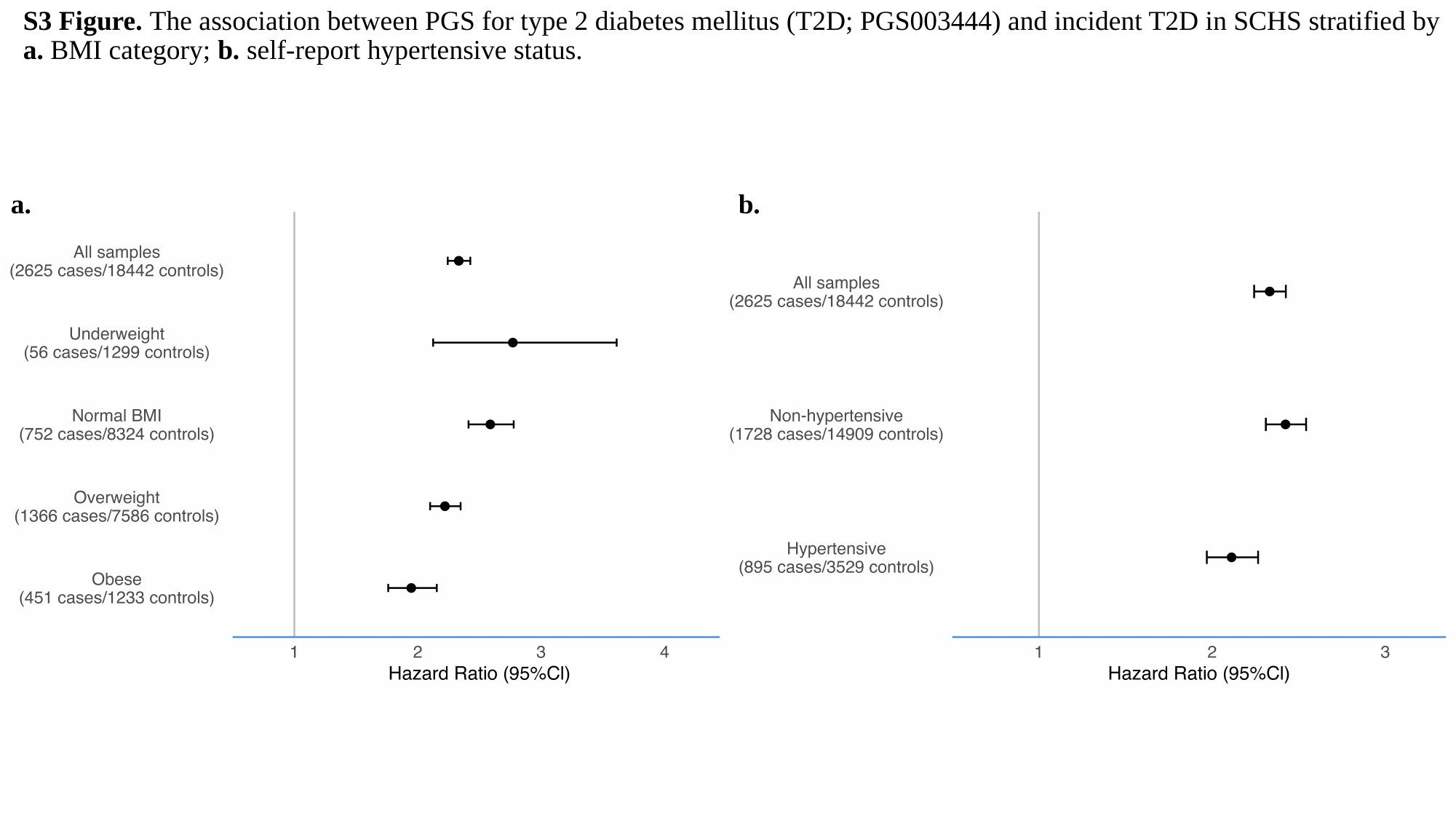

#### S3 Figure. The association between PGS for type 2 diabetes mellitus (T2D; PGS003444) and incident T2D in SCHS stratified by a. BMI category; b. self-report hypertensive status.
a.
b.
